# Analytical and clinical evaluation of antibody tests for SARS-CoV-2 serosurveillance studies used in Finland in 2020

**DOI:** 10.1101/2021.01.21.21250207

**Authors:** Nina Ekström, Camilla Virta, Anu Haveri, Timothée Dub, Lotta Hagberg, Anna Solastie, Pamela Österlund, Terhi Vihervaara, Iris Erlund, Hanna Nohynek, Merit Melin

## Abstract

**Background:** Sensitive and highly specific antibody tests are critical for detection of SARS-CoV-2 antibodies especially in populations where seroprevalence is low.

**Aim:** To set up, optimize and evaluate the analytical and clinical performance of a new in-house microsphere immunoassay for measurement of IgG antibodies to SARS-CoV-2 nucleoprotein for assessment of population seroprevalence in Finland.

**Methods:** We set up a new in-house microsphere immunoassay (FMIA) with SARS-CoV-2 nucleoprotein and optimized its analytical performance. For evaluation of clinical performance, we tested sera collected in a well-characterized cohort of PCR positive-confirmed SARS-CoV-2 patients (n=89) with mostly mild symptoms, and before the COVID-19 pandemic (n=402), for nucleoprotein specific IgG concentrations by FMIA and a commercial chemiluminescent immunoassay and for neutralizing antibodies by the microneutralization test.

**Results:** The analytical performance of FMIA was established in terms of sensitivity, linearity and precision. FMIA discriminated between COVID-19 patient and control samples with high specificity (100%) and sensitivity (100%). We generated FMIA seropositivity cut-offs, 0.46 and 1.71 U/ml, for low- and high-seroprevalence settings, respectively. In addition, we obtained high level of agreement between FMIA results and results by the microneutralization test.

**Conclusion:** The fluorescent microsphere immunoassay showed excellent analytical and clinical performance and is well suited for serosurveillance studies of SARS-CoV-2. However, to optimize analytical sensitivity and clinical specificity of the assay, different seropositivity thresholds depending on the intended use of the assay and the target population, may be needed.

## Introduction

The newly emerged SARS-CoV-2 (severe acute respiratory syndrome coronavirus 2) has resulted in worldwide pandemic. Detection of antibodies against SARS-CoV-2 is essential in population level surveillance studies and in outbreak monitoring as antibodies reveal evidence of previous infection. Antibody tests are especially important for detecting previous infections in people who had few or no symptoms. Knowledge on the spread of the virus and the protective status of the population help inform public policy makers how to manage the pandemic.

All known corona viruses share similarities in the organization and expression of their genome, encoding 16 nonstructural proteins and the four structural proteins: spike (S), envelope (E), membrane (M), and nucleocapsid (N). In most population serosurveillance studies of SARS-CoV-2 the S1 domain of the Spike glycoprotein (S), the receptor binding domain of S (RBD) and/or the viral nucleoprotein (N) have been used as antigens in antibody tests. These viral proteins have been suggested to function as major immunogenic antigens towards which most if not all those infected with SARS-CoV-2 produce antibodies. Several types of immunoassays have been described which allow detection of the presence of IgG, IgA and IgM in response to SARS-CoV-2 infection i.e. automated chemiluminescent immunoassays (CLIA), enzyme-linked immunosorbent assays (ELISA), microsphere-based immunoassays and rapid immunochromatographic tests.

A test that measures the presence of neutralizing antibodies against SARS-CoV-2 is considered the golden standard. Several studies have demonstrated that SARS-CoV-2 infection is able to induce neutralizing antibodies in most subjects and that the strength of the immune response correlates with disease severity [1-4]. However, it is possible that COVID-19 patients who experience mild symptoms may not have detectable levels of neutralizing antibodies [5]. The development of humoral immunity in COVID-19 patients seems to take a median of two weeks after onset of symptoms [6-8]. While neutralizing antibodies mainly target the S protein, antibodies to nucleoprotein could serve as a sensitive measure of past infection, because nucleoprotein is produced most abundantly during the viral replication. Most COVID-19 vaccines and vaccine candidates target the S protein. An antibody test based on nucleoprotein would enable differentiation between immunity induced by infection and vaccination and could thus be used to monitor the prevalence of infections also in vaccinated populations.

When the number of infections in the population is low, even a relatively accurate antibody test will yield a significant number of false positive results, which could be caused by cross-reactive antibodies induced by endemic coronaviruses or other infections. The sensitivity and specificity of the antibody tests should be carefully evaluated and taken into account not to overestimate the population seroprevalence [9].

In this study we describe evaluation of analytical and clinical performance of an in-house microsphere immunoassay (FMIA) for assessment of population seroprevalence to SARS-CoV-2 in Finland. We measured serum IgG antibodies to nucleoprotein in a well-characterized cohort of PCR positive-confirmed COVID-19 patients and population samples collected before the COVID-19 pandemic. Furthermore, we measured neutralizing antibodies by microneutralization test using SARS-CoV-2 virus cultures. We focused on serum samples collected during the late convalescent phase, because appearance of IgG antibodies in serum takes on average two weeks following SARS-CoV-2 infection. In addition, we analysed the samples by a widely used commercial automated chemiluminescence immunoassay and compared the clinical performance of the assays.

## Methods

### Serum samples

SARS-CoV-2 infection, exposure in households and production of antibodies to SARS-CoV-2 within families was investigated in Helsinki, Finland. Altogether 129 subjects from 39 households participated and gave written consent between March and June 2020. Serum samples (n=312) collected during the study were used for assessment of analytical performance of the antibody assays. For assessment of clinical performance, serum samples (n=89) from 58 subjects with PCR positive-confirmed SARS-CoV-2 infection collected during the late convalescent phase at a median of 30 days from the onset of symptoms (range 15 to 51 days) were used. The majority of the COVID-19 patients reported mild symptoms. Four patients reported severe symptoms and required hospital treatment. As a control group, serum samples collected in 2019 before the COVID-19 pandemic from 402 Finnish subjects aged 1 to 90 years of age were assessed. Demographics of the COVID-19 patients and donors for the pre -COVID-19 serum panel (residual sera from Helsinki University Hospital laboratory, HUSLAB) are described in Supplementary Tables S1 and S2.

### SARS-CoV-2 antibody assays

#### Fluorescent microsphere immunoassay (FMIA)

An in-house fluorescent microsphere immunoassay (FMIA) modified from a previously described method [10] was set up and optimized (Supplementary material). In brief, pre-COVID-19 sera diluted 1/100 and sera from COVID-19 patients diluted 1/100 and 1/1600, in-house reference serum (1/400 to 1/1 638 400) and two control sera per plate were mixed with SARS-CoV-2 nucleoprotein conjugated fluorescent microspheres. IgG antibodies were subsequently detected by R-Phycoerythrin (RPE) -conjugated secondary antibody. The plate was read on MAGPIX® system using xPONENT software v4.2 (Luminex®Corporation, Austin, TX). The in-house reference sample was assigned an arbitrary IgG antibody concentration of 100. Median fluorescent intensity (MFI) was converted to FMIA unit/ml (U/ml) by interpolation from a 5-parameter logistic (5-PL) reference curve. When serum was analysed in two dilutions, the IgG concentration was calculated for the 1/100 dilution and as the mean concentration of the two dilutions.

#### Abbott SARS-CoV-2 IgG test (Abbott test)

The quantitative analysis of SARS-CoV-2 IgG antibodies directed against the nucleoprotein was carried out using the SARS-CoV-2 IgG kit (Abbott Laboratories, IL, USA) on an ARCHITECT analyser in accordance with the manufacturer’s instructions. The ARCHITECT System calculates the calibrator mean chemiluminescent signal from three calibrator replicates, and the results are reported by dividing the sample result by the calibrator result. The default result unit for the SARS-CoV-2 IgG assay is index (S/C). For interpretation of the negative/positive sample result the cutoff index (S/C) 1.40 provided by the manufacturer was used.

#### Microneutralization test (MNT)

The cytopathic effect (CPE)-based microneutralization test (MNT) was performed as previously described [11]. SARS-CoV-2-specific neutralizing antibodies were detected from 2-fold serially diluted serum samples, starting from 1/4, using hCoV-19/Finland/1/2020 (GISAID accession ID EPI_ISL_407079) and hCoV-19/Finland/FIN-25/2020 (EPI_ISL_412971) viruses imported to Finland from China and Italy, respectively. Results were expressed as MNT titers corresponding to reciprocal of serum dilution that inhibited 50% of SARS-CoV-2 infection observed by CPE of inoculated cells. MNT titer was considered positive when ≥6 by either or both of the SARS-CoV-2 strains and borderline when 4≤ titer<6 by both strains.

### Evaluation of analytical performance

A serum panel from COVID-19 patients was used to quantify limits of detection (LOD) and quantification (LOQ), and assess linearity and precision of FMIA. LOD and LOQ (U/ml) were assessed by averaged MFI values generated from blank wells (n=104) and from this, mean + 3× and 10× standard deviations calculated, respectively, and interpolated from the 5-PL reference curve with GraphPad Prism v7. To assess linearity, FMIA was performed with five 4-fold dilutions (1/100 to 1/1600) of six sera, and the resulting log2-transformed IgG concentrations were compared by linear correlation analysis. Optimal linearity would yield Pearson correlation coefficient R^2^ ≥0.95. Additionally, linearity was assessed by comparing IgG concentrations obtained by dilutions 1/100 and 1/1600 of 312 sera.

Precision of FMIA was assessed by intra-assay (repeatability) and inter-assay variation (intermediate precision). Repeatability was determined by assaying 16 sera (range 0.04 to 5.92 FMIA U/ml) in duplicate and by assaying six reference serum dilutions in duplicate within a plate on 62 plates and between two plates analysed on the same day (n=26). The coefficient of variation (CV) between duplicate results was calculated and averaged. Intermediate precision (day to day) was assessed by seven sera in five separate runs, by comparing concentrations of six reference serum dilutions in 26 runs and of two control sera on 54 runs on different days. The CV between results obtained on different days was calculated and averaged. Intermediate precision was additionally assessed between results obtained by four different batches of conjugated microspheres, by three lots of RPE-conjugated secondary antibody and by four technicians with panels of sera (n=14-39). CV≤ 20% was considered acceptable precision.

The analytical performance of the Abbott test was evaluated by quantifying accuracy and precision. In the absence of a standard reference serum specimen accuracy was evaluated by comparing the replicate values (n=7-9) of negative and positive quality control samples, provided by the manufacturer, to the target values (S/C index) indicated by the manufacturer, i.e. 0-0.78 for the negative and 1.65-8.40 for the positive quality control sample, respectively. The precision of the Abbott test was evaluated by analyzing inter-assay variation (expressed by CV) of a negative and positive control sample provided by the manufacturer on seven runs/days, and two positive in-house control serum samples on three runs/days. CV≤ 20% was considered acceptable precision.

Analytical performance of MNT was evaluated by assessing precision. Inter-assay variation of MNT was evaluated by calculating CV of the titers of a positive in-house control serum included in each assay.

Intermediate precision of MNT was assessed by analyzing a panel of sera (n=98) at two time points five months apart and calculating mean CV of the titers.

### Evaluation and comparison of the clinical performances of FMIA, Abbott test and MNT

The ability of FMIA to discriminate PCR-positive confirmed COVID-19 patients and controls was assessed for IgG concentrations by analysis of receiver operating characteristic (ROC) curve and area under curve (AUC, 95% CI) with GraphPad Prism v7 (San Diego, USA). Specificity and sensitivity optimized cutoff was determined for IgG concentrations calculated for serum dilution 1/100 of patient and control samples. In addition, the ROC analysis was performed for patient samples using the mean of IgG concentrations derived from two serum dilutions; 1/100 and ≥ 1/1600. Serum samples from 57 patients collected ≥15 days after onset of symptoms were included in the analysis.

For assessment of clinical specificity 402 serum samples collected before the COVID-19 pandemic were analysed by each test. Assessment of clinical sensitivity was performed by analysing 73 to 88 sera (depending on the test) from PCR positive-confirmed COVID-19 patients. Sera were collected ≥15 days post onset of symptoms (median 30, range 15-51 days). Two sera, taken 16 and 32 days post onset of symptoms, from a seronegative subject were excluded from the analysis.

## Ethical statement

The investigations were carried out in accordance with the General Data Protection Regulation (Regulation (EU) 2016/679 and Directive 95/46/EC) and the Finnish Personal Data Act (Finlex 523/1999). The Finnish Communicable Diseases Act (Finlex 1227/2016) allows sampling for diagnostic and surveillance purposes.

Finnish population serum samples were collected during 2019. The study protocol was approved by the Ethics Committee of the Department of Medicine, Helsinki University Hospital (Permission 433/13/03/00/15).

## Results

### Analytical performance of the FMIA test

The analytical sensitivity of FMIA was defined by the limit of detection as 0.02 FMIA U/ml and the limit of quantitation as 0.07 FMIA U/ml. To assess linearity, the correlation coefficients obtained by linear correlation analysis ranged between 0.10 and 0.90 for IgG concentrations obtained by dilutions 1/100 to 1/1600, between 0.56 and 0.98 for dilutions 1/200 to 1/1600, and between 0.97 and 1.00 for dilutions 1/400 to 1/1600. An example displaying results of the linear correlation analysis is shown in Supplementary Figure S1. When IgG concentrations (n=312) calculated using 1/100 and ≥1/1600 diluted serum were compared, the IgG concentrations obtained with 1/100 dilution were on average 18% of the concentrations obtained with dilution ≥1/1600.

For assessment of precision, the mean CV for the intra-assay variation within a plate was 7 % and 8% between plates analysed on the same day. Inter-assay variation, determined for IgG concentrations of seven samples run on five days, and for two control samples on 54 days, was 13% and 19%, respectively. For assessment of intermediate precision the mean CV between results generated by four different batches of conjugated microspheres ranged from 12 to 20%, for three different lots of RPE-conjugated secondary antibody CV was 10 %, and between results obtained by four technicians CV ranged from 10 to 25 %. Overall, both intra- and inter-assay variation was found acceptable, and the precision of the FMIA established.

### Analytical performance of the Abbott test

Accuracy was estimated by evaluating the results of the control samples provided by the manufacturer. The S/C ratios of the negative and positive controls were all in line with the specifications provided by the manufacturer showing mean S/C ratios of 0.06 (range 0.06-0.07) and 3.34 (range 3.19-3.42) over the seven/nine replicates of the negative and positive sample, respectively. For assessment of precision, the inter-assay CVs of the negative and positive controls provided by the manufacturer were 6% and 2%, respectively. The CVs for the two in-house control samples were 2% and 3% agreeing with the acceptance criteria, and showing excellent intermediate precision of the assay. For standardizing the measurements, the laboratory took part in the External Quality Assessment Scheme organized by Labquality, Helsinki, Finland. Three samples were classified as negative/positive with 100% accuracy.

### Analytical performance of the MNT

The inter-assay variability of MNT was assessed by including an in-house control serum on each run. The CV of the serum was 42% (n=39, mean MNT titer 596) and 33% (n=27, mean MNT titer 526) for Fin/25/20 and Fin/1/20 viruses, respectively. Intermediate precision of the MNT was assessed at two time points five months apart and yielded CV=28% (Supplementary Figure S3).

### Clinical performance of the FMIA IgG antibody test

Because linearity analysis indicated that serum dilution 1/100 generates lower IgG concentrations than more diluted serum, the clinical specificity and sensitivity of FMIA were evaluated using two different IgG concentration cut-offs generated by ROC analysis. IgG concentration data using 1/100 diluted sera collected from COVID-19 patients and controls generated ROC curve with an area under curve 0.998 (95% CI 0.995-1). The ROC generated cut-off 0.46 U/ml resulted in 97.5% specificity (Wilson/Brown 95% CI 95.5-98.6). Sensitivity of FMIA was 96.0% (95% CI 80.5, 99.3), 100% (77.2, 100.0) and 100% (92.9, 100.0) for samples collected 15-21, 22-28 and >28 days after onset of symptoms, respectively. When ROC analysis was performed using IgG concentration data obtained from the mean concentration calculated from 1/100 and ≥1/1600 diluted serum from COVID-19 patients it yielded an area under curve 1.00 (95%CI 1.00, 1.00) and generated cut-off of 1.71 U/ml. This latter cut-off resulted in 100% specificity (95% CI 99.1 -100%) and 100% sensitivity for samples taken 15-21, 22-28 and >28 days after onset of symptoms (Table 1, Figure 1).

**Table 1.**
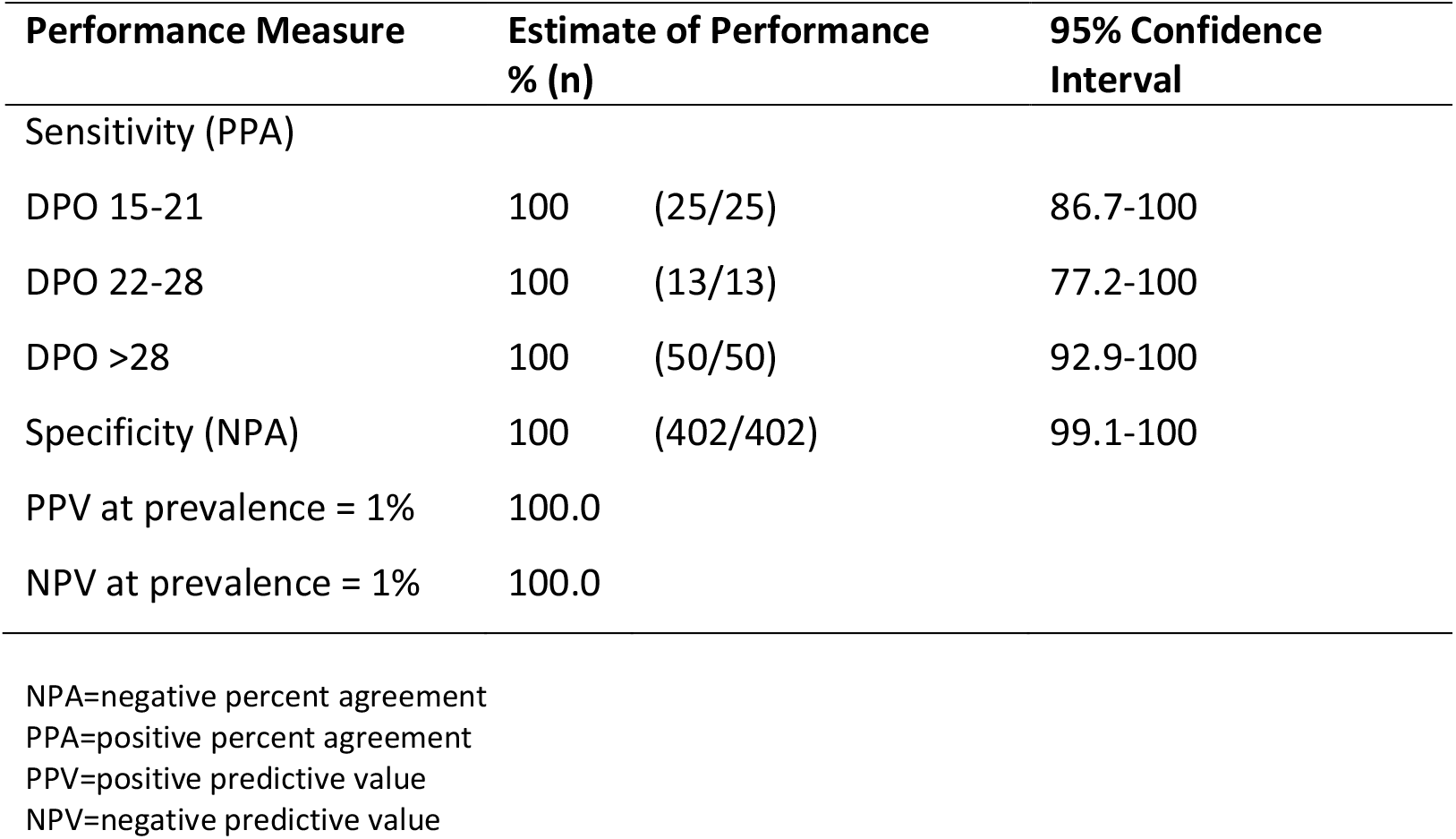
Sensitivity (NPA) and specificity (PPA) of FMIA using IgG concentration cut-off 1.71 U/ml. Sensitivity was determined according to days after onset of symptoms (DPO) at sample collection; 15-21, 22-28 and >28 days. Positive (PPV) and negative (NPV) predictive values at 1% prevalence are shown.

| Performance Measure | Estimate of Performance<br>% (n) |  | 95% Confidence<br>Interval |
| --- | --- | --- | --- |
| Sensitivity (PPA) |  |  |  |
| DPO 15-21 | 100 | (25/25) | 86.7-100 |
| DPO 22-28 | 100 | (13/13) | 77.2-100 |
| DPO >28 | 100 | (50/50) | 92.9-100 |
| Specificity (NPA) | 100 | (402/402) | 99.1-100 |
| PPV at prevalence = 1% | 100.0 |  |  |
| NPV at prevalence = 1% | 100.0 |  |  |

**Figure 1.**
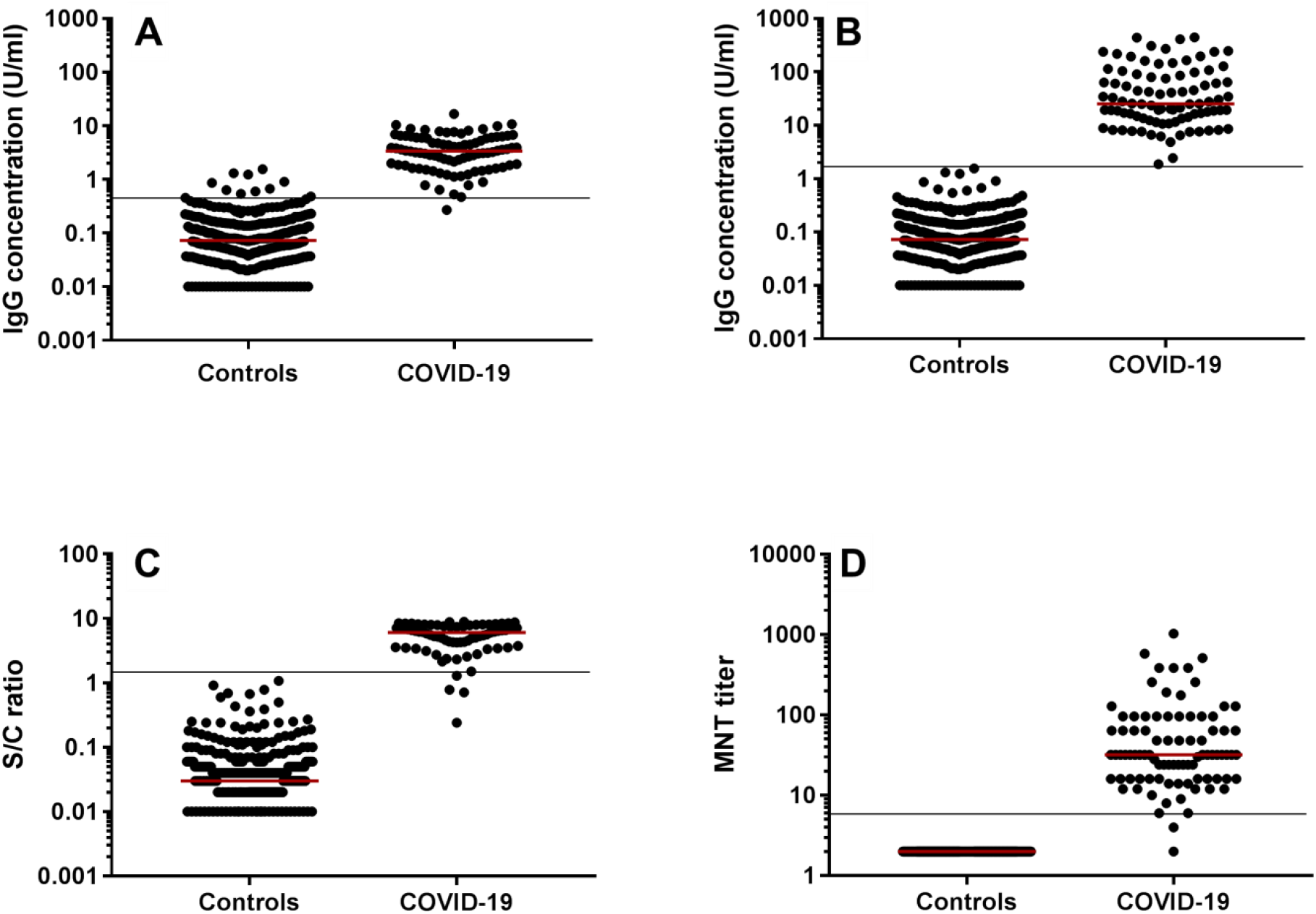
Ability of the three assays to identify COVID-19 patients. A. FMIA with cut-off 0.46 U/ml (determined by serum dilutions 1/100) B. FMIA with cut-off 1.71 U/ml (determined by mean of 1/100 and 1/1600 serum dilutions) C. Abbot IgG test and D. microneutralization test (MNT). Control sera (n=402) and sera from PCR positive-confirmed COVID-19 patients collected ≥15 days post onset of symptoms at sample collection (n= 80-88) were tested. Assay specific seropositivity cutf-offs are shown by a black line and median by a red line.

### Clinical performance the Abbott test

All sera collected before the COVID-19 pandemic were identified as true negative by the Abbott test resulting in 100 % specificity (Table 2, Figure 1). To assess sensitivity, the test identified 69/73 samples as true positive. The sensitivity was 85.7%, 100.0% and 97.6% for samples taken 15-21, 22-28 and >28 days post onset of symptoms, respectively. Four samples, taken 15, 17 (two samples) and 29 days post onset of symptoms, were identified as false negative. For two of these subjects a later sample taken 29 days post onset of symptoms was identified as true positive. For one subject both samples, taken 15 and 29 days post onset of symptoms, were identified as false negative.

**Table 2.** Sensitivity (NPA) and specificity (PPA) of the Abbott SARS-CoV-2 IgG test. Sensitivity was determined according to days after onset of symptoms (DPO) at sample collection; 15-21, 22-28 and >28 days. Positive (PPV) and negative (NPV) predictive values at 1% prevalence are shown.

| Performance Measure | Estimate of Performance<br>% (n) |  | 95% Confidence<br>Interval (%) |
| --- | --- | --- | --- |
| Sensitivity (PPA) |  |  |  |
| DPO 15-21 | 85.7 | (18/21) | 65.4-95.0 |
| DPO 22-28 | 100 | (11/11) | 74.1-100 |
| DPO >28 | 97.6 | (40/41) | 87.4-99.6 |
| Specificity (NPA) | 100 | (402/402) | 99.1-100 |
| PPV at prevalence = 1% | 100.0 |  |  |
| NPV at prevalence = 1% | 99.9 |  |  |

NPA=negative percent agreement PPA=positive percent agreement PPV=positive predictive value NPV=negative predictive value

### Performance of the MNT test

All sera collected before the COVID-19 pandemic were identified as true negative by the MNT resulting in 100 % specificity (Table 3, Figure 1). The sensitivity of MNT was 96.0%, 100% and 100.0% for samples taken 15-21, 22-28 and >28 days post onset of symptoms at sample collection (Table 3). The SARS-CoV-2 strains imported from China and Italy identified 98% and 94% of samples as true positive, respectively, and both strains identified 93% of the samples as true positive. One sample, taken 16 days post onset of symptoms, was identified as false negative by both strains, but had a borderline titer 4 to 6.

**Table 3.** Clinical sensitivity (NPA) and specificity (PPA) of the MN test using titer ≥6 as the cut-off. Sensitivity was determined according to days post onset of symptoms (DPO) at sample collection; 15-21, 22-28 and >28 days. Positive (PPV) and negative (NPV) predictive values at 1% prevalence are shown.

| Performance Measure | Estimate of Performance<br>% (n) |  | 95% Confidence<br>Interval (%) |
| --- | --- | --- | --- |
| Sensitivity (PPA) |  |  |  |
| DPO 15-21 | 96.0 | (24/25) | 79.7-99.9 |
| DPO 22-28 | 100.0 | (13/13) | 77.2-100.0 |
| DPO >28 | 100.0 | (51/51) | 93.3-100.0 |
| Specificity (NPA) | 100.0 | (402/402) | 99.1-100.0 |
| PPV at prevalence = 1% | 100.0 |  |  |
| NPV at prevalence = 1% | 100.0 |  |  |

### Comparison of SARS-CoV-2 antibody assays

All three assays identified each serum taken before the COVID-19 pandemic as true negative resulting in equal specificity of the assays. However, sensitivity of the assays varied by timing of sample collection and between assays. Sensitivity varied most for samples taken <22 days after onset of symptoms. FMIA with cut-off 1.71 U/ml solely identified samples taken <22 days post onset of symptoms as true positive suggesting high analytical sensitivity. The results obtained by the quantitative assays, FMIA and the Abbott test, showed strongest correlation (r=0.67-0.90, *P*<0.05; Figure 2C). Furthermore, results obtained by FMIA and the Abbott test correlated modestly, but statistically significantly with MNT titers for samples taken 15 to 21, and >28 days after onset of disease (Figure 2A-B). The highest agreement between FMIA and MNT was seen for samples taken >28 days after onset of symptoms.

**Figure 2.**
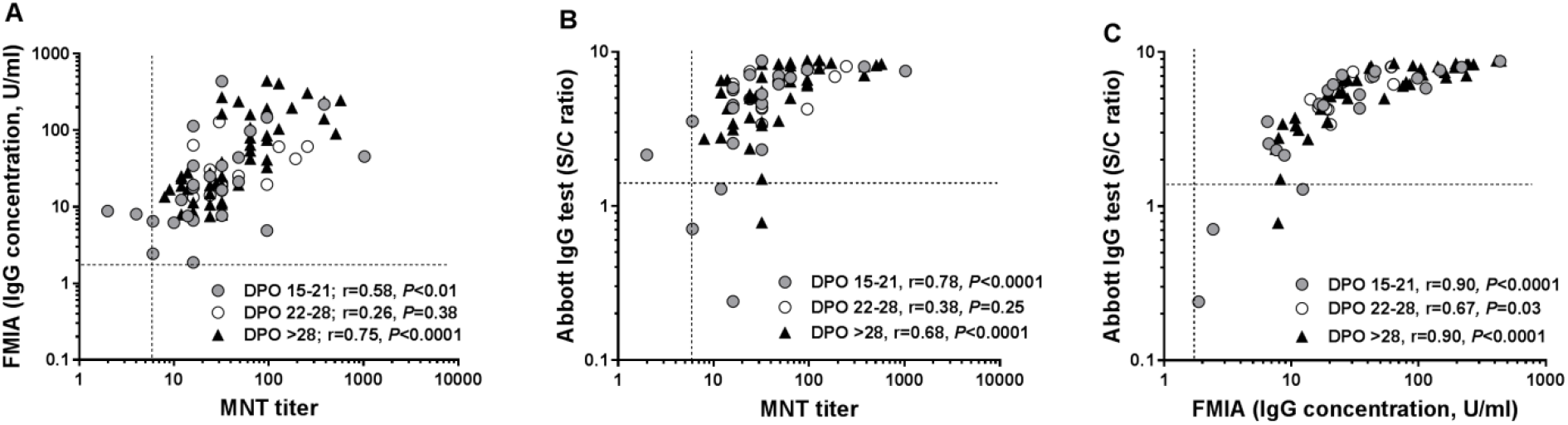
Comparison of FMIA, Abbot IgG test and microneutralization (MNT) test results with samples from PCR positive-confirmed COVID-19 patients by days post onset of symptoms at sample collection (DPO), A-C. Assay cut-off values (1.71 U/ml for FMIA, S/C ratio 1.4 for Abbot IgG test and titer 6 for MNT) are indicated with dotted lines. Spearman correlation coefficients (r) are shown.

## Discussion

Several serological assays for detection of antibodies to SARS-CoV-2 have been developed in various laboratories and are commercially available. Careful assessment of the analytical and clinical performance of the assays using clinical specimens is of great clinical importance. Large population-based seroprevalence studies are essential in providing insight into the spread of the virus and the protective status of the population. Knowledge on the performance of the assays used in seroprevalence studies is essential to provide reliable seroprevalence data. In this study, we described the development of a fluorescent microsphere immunoassay for detection of antibodies against SARS-CoV-2 nucleoprotein, used in evaluation of SARS-CoV-2 seroprevalence in Finland. The analytical performance of FMIA was established in terms of sensitivity, linearity and precision, and it was able to discriminate samples from COVID-19 patients with high specificity and sensitivity. In addition, we showed high level of consistency of the FMIA results to results obtained by the microneutralization test.

The accuracy of reporting specificity and sensitivity of an assay is greatly dependent on clinical sample selection for the evaluation. Previous reports show that milder COVID-19 disease results in lower levels of antibodies and later seroconversion than shown in severe disease [3,12,13]. The statistical outcome may not be the same when using sera of from COVID-19 patients with severe symptoms or sera from population-based serosurveillance studies where the majority of subjects will not develop severe symptoms. This increases the importance of high specificity of an assay used in population seroprevalence studies. The majority of sera from COVID-19 patients used for assessment of specificity and sensitivity in our study were from patients with mild symptoms. This is in agreement with the intended use of the test i.e. seroprevalence studies with low-disease prevalence situation.

The FMIA was developed for assessment of seropositivity in population. In the prevailing low-seroprevalence setting it was essential to optimize the analytical sensitivity and clinical specificity of the assay. This could be done using low serum dilution (1/100) and seropositivity cut-off of 0.46 U/ml, but would result in clinical sensitivity of 97.5% for samples collected 15-22 days post onset of symptoms. Thus, to optimize analytical sensitivity and clinical specificity, FMIA could be used as the first screening test and be supplemented with another assay, such as neutralization test or a second test based on spike protein antigens to exclude false positive results. By using the ROC generated higher seropositivity cut-off 1.71 U/ml, both clinical specificity and sensitivity of FMIA could be optimized to 100%. This FMIA cut-off is ideal when high seropositivity is expected. Consequently, the optimal threshold of FMIA depends on the intended use of the test and the target population. Moreover, we showed that low serum dilutions (1/100, 1/200) resulted in lower antibody concentrations than higher serum dilutions (≥1/1600). This should be taken into account when comparing antibody concentrations over time or between groups; samples should be analysed by the same serum dilutions or the concentration calculated as the mean of more than one serum dilution.

The clinical performance of the Abbott test has been assessed in various previous studies which have reported relatively high specificity, but varying sensitivity with regard to disease severity and time between symptom onset and sampling [14,15]. Using the manufacturer’s recommended index value cutoff for seropositivity we reported an assay specificity of 100% and sensitivity ≥85.7% at ≥15 days post onset of symptoms. For samples taken <22 days after onset of symptoms we reported lower sensitivity than given by the manufacturer.

In this study with majority of patients having mild disease, all three assays had sensitivities comparable to those reported for hospitalized patients (for samples taken 22 to 51 days post symptom onset). However, differences in assay sensitivity may become apparent later on when antibody levels decline. A wide range of SARS-CoV-2-neutralizing antibody titers have been reported after infection and these vary depending on the length of time from infection and the severity of disease [16]. For those who develop low neutralizing antibody response, titers have been reported to return to baseline in three to four months whereas binding antibody levels were maintained [17]. This suggests that over time the clinical sensitivity of MNT may decrease emphasizing the importance of assays measuring binding antibodies.

Limitations of this study include that the sample size, especially for assessment of sensitivity by days post onset of symptoms, was relatively small. Another limitation was that because only few patients had been hospitalized, we could not assess differences in performance that depend on disease severity. On the other hand, as the majority of the samples used in assessment of performance were from COVID-19 patients with mild symptoms, they well reflect the general population in seroprevalence studies. Furthermore, testing potentially cross-reactive sera from patients with recent infection of endemic human coronaviruses or other respiratory pathogens was not done due to lack of suitable sera.

In the future, the capacity of the FMIA-based assays can be utilized in antibody testing within SARS-CoV-2 vaccine evaluation. The flexibility of FMIA will allow several additional SARS-CoV-2 antigens to be incorporated easily into a single multiplex assay. Importantly, by measuring antibodies to the SARS-CoV-2 nucleoprotein, immune responses induced by infection and potential circulation of the virus can be assessed also in vaccinated populations.

## Data Availability

The datasets generated during and/or analysed during the current study are available from the corresponding author on reasonable request, excluding any personal data.

## Acknowledgements

We thank Hanna Valtonen, Katja Lind, Heidi Hemmilä, Tiina Pulliainen, Marja-Liisa Ollonen, Ira Greinert, Elina Järvensivu, Leena Saarinen and Marja Suorsa for their expert technical assistance and Esa Ruokokoski for data management. Christel Pussinen, Minna Haanpää, Katri Keino, Riitta Santanen and Mervi Nosa are thanked for their contribution to Finnish population sera collections. We also thank all the COVID-19 patients who participated in the study.

We gratefully acknowledge the authors, originating and submitting laboratories of the sequences from GISAID’s database. All submitting laboratories may be contacted directly via the GISAID website www.gisaid.org.

